# Role of renal venous oxygen pressure for renal function monitoring after related living-donor kidney transplantation: cohort study

**DOI:** 10.1101/2023.06.15.23291421

**Authors:** Diego Escarramán-Martínez, Montserrat Fernández-Bravo, Ashuin Kammar-García, Manuel Alberto Guerrero-Gutiérrez, Héctor David Meza-Comparán, Javier Mancilla-Galindo, Jesús Salvador Sánchez-Díaz, Emilio Cendejas-Ríos, Carla Adelina Escorza-Molina, Lorena Noriega-Salas, Germán Bernaldez-Gómez

**Author notes:** Corresponding author: Diego Escarramán-Martínez MD, MSc. Departamento de Anestesiología, Centro Médico Nacional Hospital de Especialidades “La Raza”, Instituto Mexicano del Seguro Social, Mexico City, Mexico. **Authors’ surnames and initials:** Escarramán-Martínez (DE-M), Fernández-Bravo (MF-B), Kammar-García (AK-G), Guerrero-Gutiérrez (MAG-G), Meza-Comparán (HDM-C), Mancilla-Galindo (JM-G), Sánchez-Díaz (JSS-D), Cendejas-Ríos (EC-R), Escorza-Molina (CAE-M), Noriega-Salas (LN-S), Bernaldez-Gómez (GB-G). Address reprint requests to: Diego Escarramán-Martínez MD, MSc. Departamento de Anestesiología, Centro Médico Nacional Hospital de Especialidades “La Raza”, Instituto Mexicano del Seguro Social, Mexico City, Mexico.

## Abstract

**Introduction:** Monitoring of renal function after kidney transplantation (KT) is performed by measuring serum creatinine (SCr), urine volumes (UV), and glomerular filtration rate (GFR). Other methods based on oxygen metabolism such as the renal venous oxygen pressure (P_rv_O_2_) may be useful. We aimed to explore the correlation between P_rv_O_2_ and SCr, UV, and GFR five days after KT (SCr5, UV5, and GFR5, respectively).

**Methods:** We conducted a prospective cohort study in adult patients scheduled for living-donor KT. A venous blood sample was taken from the renal vein after declamping the renal artery and blood gas determinations were made. Correlation analyses between P_rv_O_2_ and SCr5, UV5, and GFR5, were done by calculating the Spearman correlation coefficient together with generalized linear models (GLM). A Spearman correlation analysis was performed between the percentage decrease in SCr (%ΔSCr) and P_rv_O_2_. A GLM was also performed to determine the association of P_rv_O_2_ with slow graft function (SGF).

**Results:** 42 patients were included; 66.7% were men, and the median age was 31 years (IQR: 27-43.5). P_rv_O_2_ was negatively correlated with SCr5 (ρ=-0.53, p=0.003), and positively correlated with GFR5 (ρ=0.49, p=0.001) and %ΔSCr (ρ=0.47, p=0.002). A higher P_rv_O_2_ was associated with an increase in GFR in univariable (β=1.24, 95%CI: 0.56-1.93, p=0.001) and multivariable (β=1.24, 95%CI: 0.53-1.94, p=0.001) analyses. No association was found between P_rv_O_2_ and SGF.

**Conclusion:** P_rv_O_2_ could be used to monitor renal function in the first 5 days following related living-donor KT given its good correlation with SCr and GFR.

## Introduction

Kidney transplantation (KT) is the treatment of choice for patients with end-stage renal disease (ESRD). Successful KT improves the quality of life, reduces mortality, and lowers costs when compared with renal replacement therapy (RRT) (i.e., peritoneal dialysis).^1, 2^ In Mexico, there are 17,101 patients on the waiting list for KT, according to the Mexican National Transplant Center (CENATRA).^3^ One way to assess kidney graft function is through quantification of serum creatinine (SCr) levels together with daily urine volumes (UV) and glomerular filtration rate (GFR) during the first days of the postoperative period, as recommended by the Kidney Disease: Improving Global Outcomes (KDIGO) guidelines for the care of kidney transplant recipients.^4^

Ischemia-reperfusion injury (IRI) occurs as a consequence of a reduction or transient cessation of blood flow to an organ, with a subsequent restoration, causing an uncoupling between oxygen supply and consumption, leading to a progressive lesion with the outcome of cell death (mainly ferroptosis and necroptosis).^5, 6^ This type of lesion can occur in the context of organ transplantation. The kidney is an organ with very tight self-regulation, which makes it sensitive to damage by IRI.^7, 8^

The intimate correlation between peripheral oxygen saturation (SpO_2_) and partial pressure of oxygen is explained by the oxygen-hemoglobin dissociation curve. Partial venous oxygen saturation (SpvO_2_) is an informative measure of perfusion and oxygen metabolism of tissues.^9^ In terms of renal perfusion, neither the partial renal vein oxygen saturation (Sp_rv_O_2_) nor the renal venous oxygen pressure (P_rv_O_2_) have been directly measured in patients undergoing KT, however, previous studies with Clarke-type O_2_ sensors have reported renal cortical oxygen pressures (PO_2_rc) ranging from 20 to 60 mmHg, external medullary pressures between 15-30 mmHg, and internal medullary pressures <15mmHg.^10^ Currently, there is technology based on near-infrared spectroscopy aiming to predict the early recovery of renal function after reperfusion in the context of KT. This variable is known as the regional partial oxygen saturation (rSO_2_).^11^

One of the short-term complications of IRI is slow graft function (SGF), which is considered an intermediate state between immediate graft function (IGF) and delayed graft function (DGF).^12^ In general, it is defined as a lower rate of reduction of SCr in the first days after KT, without the need for RRT.^13^ Other factors have been related to the prognosis of the kidney graft, such as cold and warm ischemia times.^14–16^

The objective of this study was to explore the correlation between P_rv_O_2_ —taken directly from the renal vein after declamping the renal artery— and SCr, UV, and GFR five days after KT. The secondary objective was to assess whether P_rv_O_2_ is associated with a SGF in post-transplant patients.

## Materials and methods

### Study design

A prospective cohort study was conducted in patients scheduled for related living-donor KT at the tertiary care referral center *Centro Médico Nacional “La Raza” IMSS* in Mexico City from March 2020 to March 2022. Inclusion criteria were male or female patients older than 18 years scheduled for related living-donor KT under balanced general anesthesia. Patients for whom renal venous blood gases could not be taken were excluded. All patients were followed-up daily for five days after KT in the Transplant unit for the collection of the study variables. The present study was conducted following the Declaration of Helsinki and was approved by the Institutional Ethics and Research Committee with registration number R-2020-3501-164. All patients provided their written informed consent to participate in the study.

The three primary outcomes were SCr, UV, and GFR five days after KT. GFR was estimated according to the 2021 GFR update^17, 18^. The secondary outcome was development of SGF in post-transplant patients. Patients were categorized as having SGF or not according to the definition proposed by Wang and cols.^19^ as a SCr reduction rate of less than 20% together with SCr levels greater than 1.5 mg/dL between the first and third postoperative days. The main exposure of interest was P_rv_O_2_, sampled directly from the renal vein after declamping the renal artery. Age, body mass index, and cold ischemia time were considered as the most relevant confounding variables. Other relevant confounders were not included in the models to avoid overfitting.

### Data collection and study variables

The collection of renal vein blood samples after declamping the renal artery was performed by the surgeon responsible for the KT. Even though an attempt was made to standardize the extraction time at five minutes after declamping, due to surgical considerations and with the intention of not hindering the procedure, the extraction time ranged from 5 to 15 minutes. The procedure was performed by the surgical team by venipuncture with a 1 mL sterile syringe with 0.1 mL of 1000 IU/mL sodium heparin and a 31G needle in a single attempt, obtaining 1 mL of blood. The sample was processed in less than 15 minutes after its collection by a GEM5000 premier automated blood gas analysis equipment to determine renal blood gases (pH, renal base excess [rBE], renal lactate [rLAC], and P_rv_O_2_).

The collected variables included age, sex, and body mass index (BMI). Serum creatinine was measured pre-transplant (SCr0), on the first day (SCr1), second day (SCr2), third day (SCr3), fourth day (SCr4), and on the fifth day (SCr5). Urine volumes were measured on the first day (UV1), second day (UV2), third day (UV3), fourth day (UV4), and fifth day (UV5). Glomerular filtration rate was calculated pre-transplant (GFR0), on the first day (GFR1), second day (GFR2), third day (GFR3), fourth day (GFR4), and on the fifth day (GFR5). Warm ischemia time (WIT) was measured as the seconds between the interruption of the circulation of the donated kidney until perfusion with a hypothermic preservation solution, while cold ischemia time (CIT) was measured as the minutes from kidney preservation with the solution until transplantation to the recipient. Mean arterial pressure at declamping (MAPd) was measured in mmHg. All three were measured intraoperatively.

All patients were given balanced general anesthesia. Upon arrival in the operating room, vital signs were monitored with standard monitoring: non-invasive blood pressure, pulse oximetry, five-lead electrocardiogram, and body temperature. For fluid therapy guidance, the radial artery was cannulated to monitor the stroke volume variability (SVV), or non-invasively with the plethysmographic variability index (PVi). Induction of anesthesia was performed with fentanyl at 3 mcg/kg, cisatracurium 0.1mg/kg, propofol 1–1.5mg/kg, and lidocaine 1mg/kg. Laryngoscopy and intubation were performed conventionally, and the correct position of the endotracheal tube was confirmed with capnography and auscultation. Mechanical ventilation was carried out based on a protective strategy with a tidal volume of 6–8ml/kg of ideal weight, positive end-expiratory pressure (PEEP) of 5 cmH_2_O, plateau pressure <35 cmH_2_O, and compliance pressure <15 cmH_2_O. Maintenance was achieved with sevoflurane 0.8-1 MAC. To maintain blood pressure at target levels at the time of declamping, bolus doses of 5 to 10 mg ephedrine or 8 mcg norepinephrine were used in some patients according to the anesthesiologist’s discretion. Immunosuppression was carried out according to the local protocols as dictated by the institutional procedures’ manual.^20^

### Sample size calculation

To calculate the sample size, the estimation of a correlation coefficient of 0.5 was considered, with a power of 90% and a bilateral alpha error of 5%, giving a sample size of 37 subjects. The sample size calculation was performed using the G*Power software version 3.1.9.7.

### Statistical analyses

Quantitative variables are presented as the median with interquartile range (IQR), while qualitative variables are expressed as absolute frequencies and percentages (%). For the comparison between groups, the Mann-Whitney U test was used for quantitative variables, and the Chi-square test, or Fischer’s exact test, for qualitative variables. The difference in SCr and UV values over the first 5 days was evaluated with the Friedman test, and the post hoc comparison of each day with the preoperative value was performed with Dunn’s test. Coefficients of variation of SCr, UV, and GFR were calculated during the first 5 days post-transplantation.

For the correlation analysis between P_rv_O_2_ and SCr5, UV5 and GFR5, the Spearman correlation coefficient was calculated together with the construction of generalized linear models (GLM) with identity function taking SCr5, UV5, and GFR5 as the dependent variable, and P_rv_O_2_ as an independent variable; likewise, a Spearman correlation analysis was performed between the percentage decrease in SCr (%ΔSCr) and P_rv_O_2_ to standardize the change between the variables. The percentage decrease in SCr was calculated as follows: %ΔSCr = [(SCr0 - SCr5) / SCr5] * 100

The results of the correlation analyses are presented with the Rho statistic (ρ) and the p-value, while the results of the GLM are presented with the correlation coefficient (β) and 95% confidence intervals (95% CI). Multivariable models were created adjusting the P_rv_O_2_ regression coefficient for age, BMI, and CIT for the SCr5 and UV5 models, while for the GFR5 model only CIT and BMI were adjusted because the GFR is highly correlated with age. The fit of the models was evaluated with the coefficient of determination (R^2^). The assumed collinearity was verified using the variance inflation factor (VIF).

To determine the association of P_rv_O_2_ with SGF, a GLM was performed with a binomial identity function taking SGF as the dependent variable and P_rv_O_2_ as the independent variable. The model was adjusted with the variables age, BMI, and CIT. Results are presented as risk ratio (RR) together with their 95% CI and p-value. A p-value <0.05 was considered to determine statistical significance. All analyses were performed in R software (version 2023.03.0+386), and all figures were created in GraphPad (version 9.0.1).

## Results

Of 46 eligible patients, 42 patients were included in the analysis (4 patients were eliminated as renal venous blood gases analysis was not possible due to difficulties in the surgical technique). Most of the patients were men (n=29, 66.7%) with a median age of 31 years (IQR: 27-43.5); while the median P_rv_O_2_ was 59 mmHg (IQR: 55.7-63.0). The rest of the demographic and clinical characteristics of the patients are presented in **Table 1**. **Figure 1** summarizes the changes in SCr, UV5, and GFR. During the first 5 days after the KT, a significant decrease in the three variables was observed over time (all p<0.0001). The SCr0 was 13.6 mg/dL (IQR: 11.0-19.9) and decreased significantly on day 5 (3.5 mg/dL, IQR: 3.0-5.9) (p<0.0001). Likewise, the UV decreased significantly from the first-day post-transplantation to day 5 (5904 mL, IQR: 4387.5-6604.5 vs. 3948.5 mL, IQR: 3504-4270; p<0.0001), while GFR increased from pre-transplantation to day 5 (4 ml/min/1,73 m^2^, IQR: 1.8-11.5, vs 18.8 ml/min/1,73 m^2^, IQR 3.6-69.8; p<0.001). The mean coefficient of variation for SCr during the 5 days after transplantation was 28.1% (95% CI: 23.4-32.8); for UV, it was 17.8% (95% CI: 14.9-20.7), and for the GFR, it was 46.9% (95% CI: 40.1-53.7). **Figure 2** shows the results of the correlation analysis of P_rv_O_2_ with SCr5, UV5, and GFR5. It was observed that P_rv_O_2_ correlates negatively with SCr5 (ρ=-0.53, p=0.003), and positively with GFR5 (ρ=0.49, p=0.001) and %ΔSCr (ρ=0.47, p=0.002), but there was no correlation with the UV5 (ρ=0.09, p=0.53).

**Figure 1.**
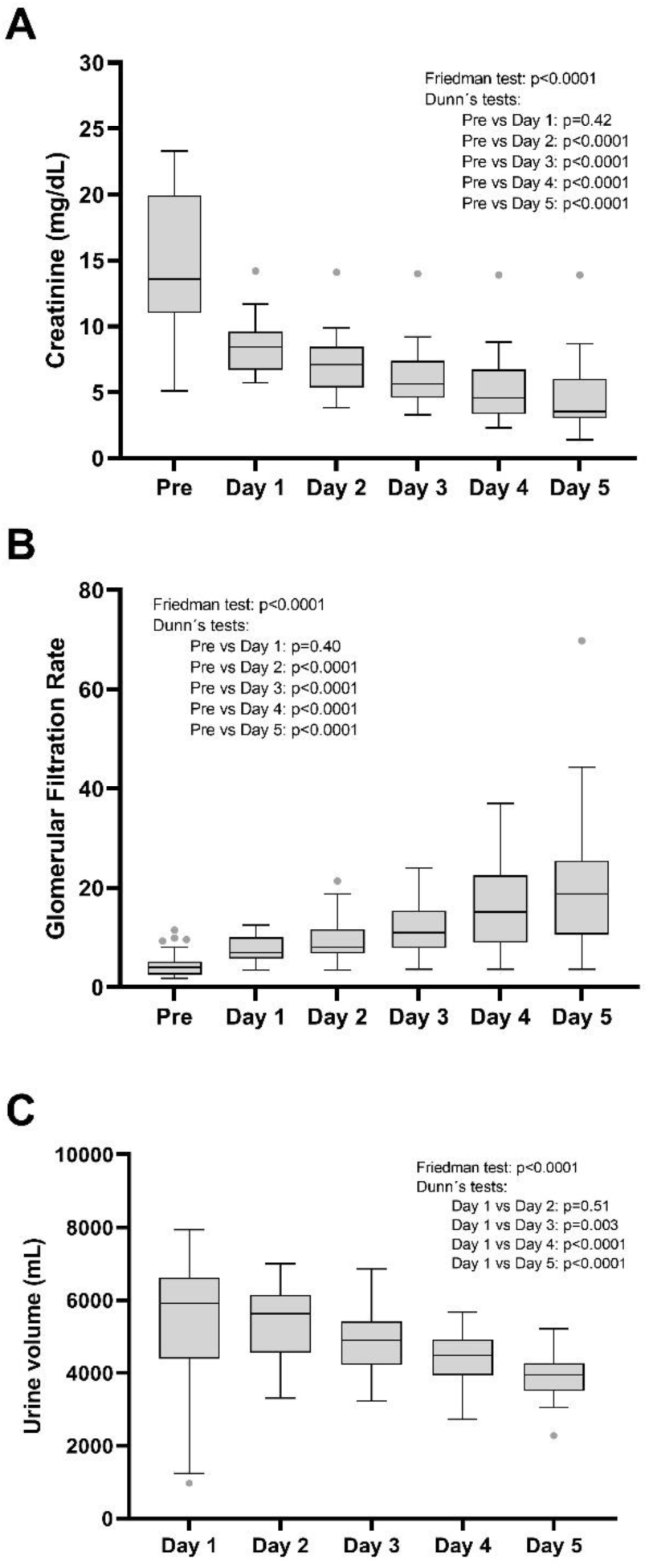
Comparisons of serum creatinine, glomerular filtration rate, and urine volume in the first 5 days post-transplant. A: Serum creatinine, B: Glomerular filtration rate, C: Urine volume. Data are presented as median, 1st quartile, 3rd quartile, and Tukey hinges. Data were compared by the Friedman test. The post hoc comparison was made by Dunn’s test.

**Figure 2:**
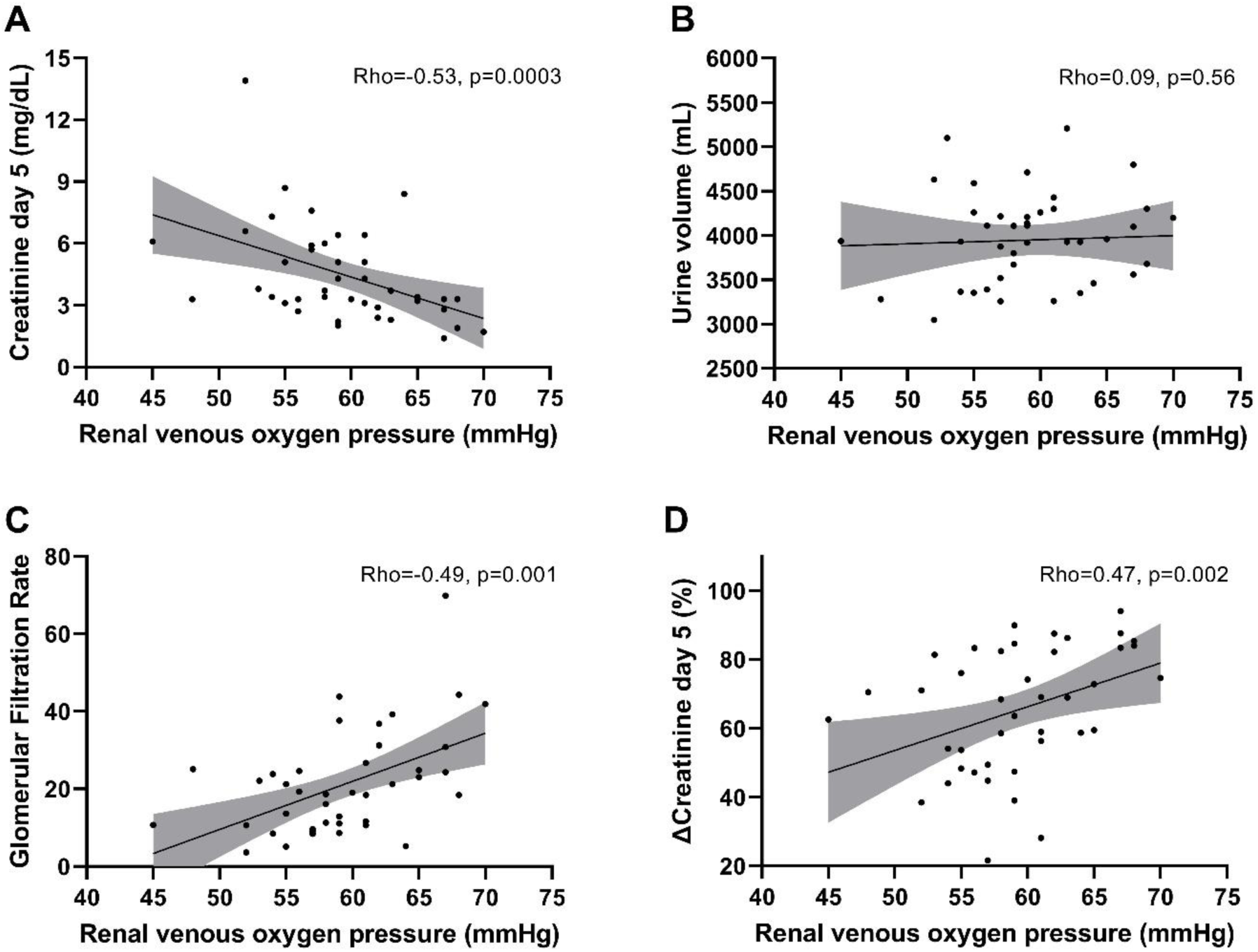
Correlation analysis between renal venous oxygen pressure and serum creatinine levels and urine volume on day 5 post-transplant. A: Serum creatinine, B: Urine volume, C: Glomerular filtration rate, D: Percentage decrease in serum creatinine. The correlation slope and 95% CI are presented. The analyses were performed by Spearman’s correlation test.

**Table 1.** Demographic and clinical data of the study sample

| Table 1. Demographic and clinical data of the study sample |  |
| --- | --- |
|  | Total<br>n=42 |
| **Demographic data** |  |
| Sex, (%) |  |
| Female | 14 (33.3) |
| Male | 29 (66.7) |
| Age, years | 31 (27 – 43.5) |
| Body mass index | 23.8 (23 – 25.1) |
| **Clinical data** |  |
| Mean arterial pressure at declamping, mmHg | 93 (88 – 99) |
| Cold ischemia time, minutes | 59 (56.7 – 70.5) |
| Warm ischemia time, seconds | 164.5 (131.5 – 226.7) |
| Renal pH | 7.31 (7.28 – 7.32) |
| Renal base excess, mg/dL | -4.8 (-5.4 – -3.9) |
| Renal lactate, mg/dL | 1.45 (1.0 – 1.6) |
| Renal venous oxygen pressure, mmHg | 59 (55.7 – 63.0) |
| Data are presented as median (IQR) or frequency (%) |  |

**Table 2** shows the results of the GLM, where the inversely proportional relationship of P_rv_O_2_ with SCr was observed (β=-0.202; 95% CI: -0.328 to -0.075; p=0.002). These results were maintained after adjusting for age, BMI, and CIT (β=-0.172; 95% CI: -0.300 to -0.043; p=0.01, R^2^=0.32), however, there was no significant relationship for UV. On the other hand, GFR is affected by P_rv_O_2_, since it was observed that as P_rv_O_2_ increases, also does GFR (β=1.24, 95%CI: 0.56 to 1.93, p=0.001), even after multivariable adjustment (β=1.24, 95%CI: 0.53-1.94, p=0.001). Regarding %ΔSCr, univariable GLM shows a positive relationship with P_rv_O_2_ (β=1.271; 95%CI: 0.292 to 2.249; p=0.01), but the relationship decreases after adjusting for age, BMI, and CIT (β=0.934; 95% CI: -0.081 to 1.950; p=0.07, R^2^=0.24).

**Table 2.** Results of the generalized linear models to determine the relationship of P_rv_O_2_ with serum creatinine, GFR, and urine volume on the fifth day

|  |  |  |  |  |
| --- | --- | --- | --- | --- |
|  | Univariable model |  | Multivariable model |  |
|  | **β (95% CI)** | **p-value** | **β (95% CI)** | **p-value** |
| **Model for SCr5†** |  |  |  |  |
| P<sub>rv</sub>O<sub>2</sub> | -0.202 (-0.328 to -0.075) | 0.002 | -0.172 (-0.300 to -0.043) | 0.01 |
| Age | 0.077 (0.019 to 0.135) | 0.01 | 0.043 (-0.017 to 0.104) | 0.16 |
| BMI | 0.178 (-0.261 to 0.617) | 0.41 | 0.122 (-0.282 to 0.528) | 0.54 |
| CIT | 0.066 (-0.012 to 0.144) | 0.09 | 0.047 (-0.024 to 0.118) | 0.19 |
| **Model for UV5‡** |  |  |  |  |
| P<sub>rv</sub>O<sub>2</sub> | 4.60 (-28.74 to 37.95) | 0.78 | -5.6 (-39.47 to 28.25) | 0.73 |
| Age | -17.43 (-31.16 to -3.70) | 0.01 | -17.45 (-33.50 to -1.39) | 0.03 |
| BMI | -59 (-161.60 to 43.43) | 0.25 | -21.1 (-127 to 85.59) | 0.69 |
| CIT | -3.06 (-22.5 to 16.016) | 0.74 | 1.43 (-17.37 to 20.25) | 0.87 |
| **Model for GFR§** |  |  |  |  |
| P<sub>rv</sub>O<sub>2</sub> | 1.24 (0.56 to 1.93) | 0.001 | 1.24 (0.53 to 1.94) | 0.001 |
| BMI | -0.16 (-2.63 to 2.31) | 0.89 | -0.43 (-2.61 to 1.76) | 0.70 |
| CIT | -0.17 (-0.62 to 0.27) | 0.42 | -0.13 (-0.53 to 0.28) | 0.53 |
| **Model for %ΔSCr¶** |  |  |  |  |
| P<sub>rv</sub>O<sub>2</sub> | 1.27 (0.29 to 2.25) | 0.01 | 0.93 (-0.08 to 1.95) | 0.07 |
| Age | -0.56 (-0.99 to -0.13) | 0.01 | -0.485 (-0.97 to 0.00) | 0.05 |
| BMI | 0.33 (-2.98 to 3.64) | 0.84 | 1.15 (-2.05 to 4.35) | 0.47 |
| CIT | -0.24 (-0.84 to 0.37) | 0.43 | -0.07 (-0.63 to 0.49) | 0.81 |
| SCr5: Serum creatinine at day 5, UV5: Urine volume at day 5, P<sub>rv</sub>O<sub>2</sub>: Renal venous oxygen pressure, BMI: Body mass index, CIT: Cold ischemia time, GFR: Glomerular filtration rate.<br>†: R<sup>2</sup> 0.32, ‡: R<sup>2</sup> 0.18, §: R<sup>2</sup> 0.26 ¶: R<sup>2</sup> 0.24<br>Data are presented as regression coefficient (β) and 95% confidence interval (95% CI). |  |  |  |  |

Of the total sample, 17 patients (40.4%) developed SGF. **Supplementary Table 1** shows the comparisons of the characteristics between the patients with and without SGF, noting that the subjects with SGF had a lower SCr2 (8mg/dL, IQR: 6.2-8.9 vs 6.8mg/dL, IQR: 4.9-8; p= 0.03), SCr3 (7.2mg/dL, IQR: 5.7-8.5 vs 4.6mg/dL, IQR: 4.1-6.2; p= 0.001), SCr4 (6.7mg/dL, IQR: 5.2-8.2 vs 3.6mg/dL, IQR: 3-5.3; p= 0.001) and SCr5 (5.9mg/dL, IQR: 3.8-7.4 vs 3.2mg/dL, IQR: 2.3-3.7; p=0.001). The UV5 was similar between both groups, but there were differences in SCr from the second day to the fifth day, even though the P_rv_O_2_ was similar between both groups (58 mmHg, IQR: 55-61 vs. 60 mmHg, IQR: 56-66, p= 0.13). The results of the GLM between P_rv_O_2_ and the SGF are shown in **Supplementary Table 2**, and it was observed that there was no association between the P_rv_O_2_ and SGF.

## Discussion

The objective of this study was to explore the existing correlation between P_rv_O_2_ and SCr, UV, and GFR levels five days after KT. Our results show that P_rv_O_2_ is correlated with SCr and GFR, showing that as P_rv_O_2_ increases, SCr5 decreases, while GFR increases. The secondary objective was to evaluate if P_rv_O_2_ is associated with SGF in post-transplant patients. Our results suggest that there is no association between P_rv_O_2_ and SGF. However, likely, our study did not have sufficient statistical power to assess this secondary objective since the sample size calculation was performed for primary outcomes only.

To the best of our knowledge, this is the first study in which P_rv_O_2_ was directly quantified in post-operative patients with living-donor kidney grafts. In our study, we obtained a median P_rv_O_2_ of 59 mmHg, a result very similar to that reported in other studies with animal models.^10, 21^ Welch et al.^22^ reported a mean partial pressure of oxygen of 42 mmHg, obtained through gas analysis, while Palm et al.^23^ reported 50 mmHg, obtained by O_2_ sensors. Lastly, O’Neill et al.^24^ also reported a pressure of roughly 45 mmHg using O_2_ sensors. All these values were obtained from the renal cortex. The variability of these results can be explained by the lack of standardization of the technique used: different animal models and different measurement methods.

Patients with lower transplant SCr5 values also had higher P_rv_O_2_. Currently, renal function monitoring after renal transplantation is performed with the measurement of SCr, due to its availability and low cost. Its levels are determined both by the metabolic demand of the recipient and by the supply of nephrons from the donor.^25^ In other words, kidney function is largely determined by the body’s metabolic demand.^26^ This study suggests that P_rv_O_2_ could serve as a monitoring tool in the first 5 days after KT. At present, non-invasive methods have been proposed to estimate renal oxygen dynamics, such as near-infrared spectroscopy (NIRS), a non-invasive optical method that continuously calculates the difference between oxygenated and deoxygenated hemoglobin in a given area of tissue, thereby obtaining the rSO_2_. Vidal et al.^27^ suggested that renal rSO_2_ correlates with and predicts the function of the renal allograft in pediatric patients with an average basal rSO_2_ of 68.8%, and this, as in our study, does not correlate with the uresis, a phenomenon that can be explained by the intervention of different variables on the determinants of uresis and urine volume (consumption of oral fluids, intravenous solutions, personalized mean arterial pressures, previous fluid overload of the recipient), which is consistent with the results in our work where P_rv_O_2_ was associated with SCr5 levels but not with UV5. More recently, the pilot study by Lau et al.^11^ demonstrated the feasibility of monitoring rSO_2_ in renal allograft transplantation during the reperfusion phase, reporting an adequate correlation between rSO_2_ and allograft function up to day 14. An explanation for this is that the venous oxygen pressure is the amount of oxygen dissolved in a venous blood sample and it is proportional to the oxygen content per gram of hemoglobin, therefore, the factors that can alter the renal venous oxygen concentration are determined by the perfusion pressure, the blood volume that enters and the oxygenation of the blood that comes from the bloodstream, in such a way that the increase in the partial pressure of oxygen in venous blood after reperfusion following vascular declamping could be attributed to both an adequate supply as well as optimal microcirculatory conditions for its extraction. Hence, the deterioration of microvascular perfusion has been attributed to a reduction in the volume of total blood flow and the diameter of the endothelial glycocalyx in the peritubular capillary network.^28^ In other words, P_rv_O_2_ can be an indirect indicator of renal oxygen metabolism,^8^ thus helping to understand graft IRI.

Our study has some limitations. First, as mentioned above, it was not possible to standardize the blood gases sampling time after declamping. Second, the comparisons that were made in terms of P_rv_O_2_ results with other studies (for instance, rSO_2_, partial pressures of oxygen in the renal cortex, etc.) must be taken with caution since although all these parameters have been used to monitor renal function and are based on the metabolism of renal oxygen, strictly speaking, they are all different parameters. Additionally, regression models were adjusted for the most relevant confounders as defined by consensus between authors and were limited to three to avoid overfitting, meaning that other relevant confounders may have been left out. Likewise, the lack of association between P_rv_O_2_ and SGF could be due to insufficient statistical power to evaluate this secondary objective, so new studies should be conducted to assess this hypothesis with an adequate sample size calculation.

The main strength of our study is that it establishes the role of P_rv_O_2_ obtained directly from the renal vein after declamping with monitoring of the renal function in the context of KT. Our results contribute to a better understanding of renal metabolism and establish P_rv_O_2_ as a low-cost biomarker to monitor renal function after KT that is worth studying to further determine its prognostic value.

## Conclusion

P_rv_O_2_ may be used to monitor renal function in the first 5 days after a related living-donor KT given its good correlation, which is inversely proportional to SCr, and directly proportional to GFR. More research is needed to elucidate its prognostic value.

## Authors contributions

DE-M and MF-B: Conceptualization, Methodology, Writing - Original Draft; AK-G: Formal analysis, Visualization, Validation; MAG-G: Investigation; HDM-C: Investigation, Writing - Review & Editing; JM-G: Writing - Review & Editing, Formal analysis, Visualization, Validation; JSS-D: Supervision, EC-R, CAE-M, LN-S and GB-G: Investigation, Supervision.

## Data Availability

The data that support the findings of this study are openly available in the Harvard Dataverse Repository at https://doi.org/10.7910/DVN/VBCKWO

## Acknowledgements

None.

## List of abbreviations

BMI: Body mass index
CENATRA: Mexican National Transplant Center
CIT: Cold ischemia time
DGF: Delayed graft function
ESRD: End-stage renal disease
GFR: Glomerular filtration rate
GFR0: Pre-transplant glomerular filtration rate
GFR1: First day post-transplant glomerular filtration rate
GFR2: Second day post-transplant glomerular filtration rate
GFR3: Third day post-transplant glomerular filtration rate
GFR4: Fourth day post-transplant glomerular filtration rate
GFR5: Fifth day post-transplant glomerular filtration rate
GLM: Generalized linear model
IGF: Immediate graft function
IQR: Interquartile range
IRI: Ischemia-reperfusion injury
KDIGO: Kidney Disease Improving Global Outcomes
KT: Kidney transplantation
MAC: Minimum alveolar concentration
MAPd: Mean arterial pressure at declamping
NIRS: Near-infrared spectroscopy
PEEP: Positive end-expiratory pressure
PO_2_rc: Renal cortical oxygen pressure
P_rv_O_2_: Renal venous oxygen pressure
PVi: Plethysmographic variability index
rBE: Renal base excess
rLAC: Renal lactate
RR: Risk ratio
RRT: Renal replacement therapy
rSO_2_: Regional partial oxygen saturation
SCr: Serum creatinine
SCr0: Pre-transplant serum creatinine
SCr1: First day post-transplant serum creatinine
SCr2: Second day post-transplant serum creatinine
SCr3: Third day post-transplant serum creatinine
SCr4: Fourth day post-transplant serum creatinine
SCr5: Fifth day post-transplant serum creatinine
SGF: Slow graft function
SpO_2_: Peripheral oxygen saturation
Sp_rv_O_2_: Partial renal vein oxygen saturation
SpvO_2_: Partial venous oxygen saturation
SVV: Stroke volume variability
UV: Urine volume
UV0: Pre-transplant urine volume
UV1: First day post-transplant urine volume
UV2: Second day post-transplant urine volume
UV3: Third day post-transplant urine volume
UV4: Fourth day post-transplant urine volume
UV5: Fifth day post-transplant urine volume
VIF: Variance inflation factor
WIT: Warm ischemia time

