## Supplementary material for "Role of renal venous oxygen pressure for renal function monitoring after related living-donor kidney transplantation: cohort study"

| **Supplementary Table 1. Comparison of demographic and clinical data between patients with and without slow graft function** | | | |
| --- | --- | --- | --- |
| **Demographic data** | **SGF (n=17)** | **NSGF (n=25)** | **p-value** |
| Sex, (%) |  |  | 0.12 |
| Female | 8 (47.1) | 6 (24) |  |
| Male | 9 (52.9) | 19 (76) |  |
| Age, years | 33 (26 – 49.5) | 31 (27 – 40.5) | 0.43 |
| Body mass index | 24 (22.9 – 25.2) | 23.6 (23 – 25.1) | 0.96 |
| **Clinical data** | | | |
| Mean arterial pressure at declamping, mmHg | 91 (87.8 – 98) | 95 (89 – 99.1) | 0.41 |
| Cold ischemia time, minutes | 60 (56.5 – 76.5) | 59 (56.5 – 65.5) | 0.78 |
| Warm ischemia time, seconds | 170 (131 – 601) | 161 (126.5 – 192) | 0.17 |
| Renal pH | 7.31 (7.29 – 7.32) | 7.31 (7.27 – 7.32) | 0.74 |
| Renal base excess, mg/dL | -4.8 (-5.3 – -3.8) | -4.8 (-5.8 – -3.9) | 0.83 |
| Renal lactate, mg/dL | 1.5 (1 – 1.6) | 1.4 (0.95 – 1.6) | 0.64 |
| Renal venous oxygen pressure, mmHg | 58 (55 – 61) | 60 (56 – 66) | 0.13 |
| **Serum creatinine** | | | |
| Pre-transplantation | 13 (10.8 – 19.1) | 14.1 (10.5 – 20.2) | 0.51 |
| Day 1 | 8.7 (7.1 – 9.7) | 8.1 (6.3 – 9.4) | 0.41 |
| Day 2 | 8 (6.2 – 8.9) | 6.8 (4.9 – 8) | 0.03 |
| Day 3 | 7.2 (5.7 – 8.5) | 4.6 (4.1 – 6.2) | 0.001 |
| Day 4 | 6.7 (5.2 – 8.2) | 3.6 (3 – 5.3) | 0.001 |
| Day 5 | 5.9 (3.8 – 7.4) | 3.2 (2.3 – 3.7) | 0.001 |
| **Urine volume** | | | |
| Day 1 | 5630 (4704 – 6171) | 6100 (4350 – 7017) | 0.36 |
| Day 2 | 5208 (4475.5 – 5706) | 5930 (4764.5 – 6413.5) | 0.06 |
| Day 3 | 4982 (3725.5 – 5363.5) | 4821 (4321 – 5699) | 0.45 |
| Day 4 | 4480 (3963 – 4608) | 4500 (3922 – 5195) | 0.62 |
| Day 5 | 4108 (3567 – 4300) | 3928 (3454 – 4230) | 0.55 |
| **Glomerular filtration rate** | | | |
| Pre-transplantation | 4.3 (2.3 – 5) | 3.8 (2.7 – 5.4) | 0.91 |
| Day 1 | 6.6 (5.1 – 8.3) | 7.6 (6 – 10.9) | 0.10 |
| Day 2 | 7.5 (5.7 – 10) | 8.7 (7.7 – 14.9) | 0.007 |
| Day 3 | 8.1 (6.1 – 10.3) | 13.4 (10.6 – 18.1) | 0.001 |
| Day 4 | 8.6 (7 – 13.5) | 19.3 (13.4 – 23.7) | 0.001 |
| Day 5 | 10.6 (8.4 – 18.5) | 23.8 (17.2 – 37.2) | 0.001 |
| Data are presented as median (IQR) or frequency (%)  Comparisons were made by the Mann-Whitney U test or the Chi-square test. | | | |

| **Supplementary Table 2. Results of the generalized linear model to determine the risk of slow graft function in post-transplant patients** | | | | | | |
| --- | --- | --- | --- | --- | --- | --- |
|  | **Univariate model** | | | **Adjusted model** | | |
|  | β | RR (95% CI) | p-value | β | RR (95% CI) | p-value |
| P_rv_O_2_ | -0.07 | 0.93 (0.82 – 1.05) | 0.21 | -0.06 | 0.94 (0.83 – 1.07) | 0.35 |
| Age | 0.03 | 1.03 (0.98 – 1.09) | 0.25 | 0.02 | 1.02 (0.96 – 1.08) | 0.47 |
| BMI | 0.01 | 1.01 (0.71 – 1.45) | 0.95 | -0.02 | 0.98 (0.67 – 1.44) | 0.98 |
| CIT | 0.03 | 1.03 (0.96 – 1.10) | 0.45 | 0.01 | 1.01 (0.95 – 1.09) | 0.61 |
| P_rv_O_2_: Renal venous oxygen pressure, BMI: Body mass index, CIT: Cold ischemia time.  Data are presented as regression coefficient (β), risk ratio (RR), and 95% confidence interval (95% CI). | | | | | | |
